# Baseline ferritin predicts myocardial iron uptake following intravenous iron therapy- a hypothesis generating study

**DOI:** 10.1101/2025.01.11.25320372

**Authors:** Julio Nunez, Anna Mollar, Mayra Vera-Aviles, Syeeda Kabir, Akshay Shah, Paolo Polzella, Michael Desborough, Ingrid Cardells, Gema Miñana, Irene del Canto, Vanessa Ferreira, Stefan Piechnik, Alicia Maceira, Samira Lakhal-Littleton

## Abstract

**Aims:** Iron deficiency (ID) is a common co-morbidity in heart failure (HF). Intravenous iron therapy improves outcomes. Several mechanisms have been proposed, including myocardial iron repletion. However, it is unknown if clinical iron markers predict the extent of this repletion. The aim of this study is to address this question by harnessing data from two clinical studies that evaluated changes in myocardial iron using cardiac magnetic resonance (CMR).

**Methods and Results:** We performed a post-hoc analysis of two previously published trials. The Myocardial-IRON trial measured change in myocardial iron, denoted by a decrease in CMR T1-mapping, at 7 and 30 days after FCM in patients with ID and HF. The STUDY trial measured myocardial and spleen iron at multiple timepoints after FCM in patients with ID without HF. In this post-hoc analysis, we examined the association between baseline iron markers (Tsat and ferritin) and change in myocardial iron in the weeks after FCM therapy. Changes in spleen iron were also examined, due its role as an intermediary in the redistribution of iron from iron-carbohydrate complexes such as FCM. In patients with or without HF, higher plasma ferritin at baseline predicted lower rise in myocardial iron in the weeks after therapy with FCM. In contrast, higher plasma ferritin at baseline predicted a greater rise in spleen iron.

**Conclusions:** These data point towards the hypothesis that functional ID, which is characterized by elevated ferritin, could limit myocardial iron repletion after IV iron therapy, by favoring iron trapping in the spleen.

## INTRODUCTION

Myocardial iron uptake has been proposed as one of the mechanisms underlying the benefits of intravenous iron therapy in patients with iron deficiency (ID) and heart failure (HF). However, it is unknown whether myocardial iron uptake relates to baseline iron markers. Myocardial iron uptake can be determined non-invasively as decreased T1-mapping cardiac magnetic resonance (CMR). Using this approach, the Myocardial-IRON trial previously showed that treatment with ferric carboxymaltose (FCM) in patients with HF significantly decreased myocardial T1 at 7 days, together with improvement in global longitudinal strain at 30 days (1,2). More recently, a pharmacokinetic trial (STUDY) in patients with ID without HF demonstrated that FCM decreased myocardial T1 and this decrease was sustained at 42 days (3). STUDY also reported a marked decrease in spleen T1 at 14 days, which was partially reversed at 42 days, confirming the known role for spleen macrophages in the uptake and subsequent redistribution of iron from iron-carbohydrate complexes such as FCM.

## AIM

To examine the relationship between baseline iron markers and myocardial iron uptake after intravenous iron therapy with FCM, and the role of the spleen in this context. Understanding this relationship could help inform the ongoing debate over what markers best predict favorable response to intravenous iron therapy.

## METHODS

### Study population and procedures

These are post hoc analyses of Myocardial-IRON and STUDY trials (1-3). Details of study populations were as reported previously (1-3).

*Myocardial-IRON trial* was a multicenter, randomized, double-blind, placebo-controlled trial (NCT03398681) to assess the effect of intravenous FCM versus placebo on myocardial iron repletion at 7 and 30 days in patients with HF and ID. Detailed information about the study were published elsewhere (1,2) This study is registered at http://clinicaltrials.gov (NCT03398681).

The <u>S</u>tudy of <u>T</u>issue Iron <u>U</u>ptake in patients iron <u>D</u>eficienc<u>Y</u> (STUDY) was an investigator-initiated, single-centre, prospective, observational study (NCT05609318). It included 12 patients without HF receiving standard of care FCM for the correction of ID. Detailed information and main results were published elsewhere (3). The study was registered prospectively on the ISRCTN registry (ISRCTN15770553) and ClinicalTrials.gov (NCT05609318).

### Statistical analysis

In Myocardial-IRON trial, the relationships between baseline ferritin and TSAT and changes in T1-mapping, and Δ3D-GLS were assessed using linear mixed-effect models, adjusted for baseline value of the endpoints (ANCOVA design), age, sex, and participant sites. In the STUDY cohort, within-group unadjusted comparisons using linear regression was used to examine the association between baseline ferritin and TSAT and changes in myocardial and spleen T1-mapping at 42 days. Estimates were presented as least square means with 95% CIs. A two-sided p-value<0.05 was considered significant, and no adjustments were made for multiple comparisons. All analyses were performed using STATA 18.0 (Stata Statistical Software, College Station, TX).

## RESULTS

In Myocardial-IRON trial, the median age of the sample was 73 years, with 75.5% men, and 94.3% patients in New York Association class II. Median NT-proBNP was 1690 pg/mL, and all patients fulfilled ESC ID criteria. The median (Q1 to Q3) ferritin and TSAT were 63 ug/L (33-114) and 15.7% (11-19.2), respectively. Baseline mean Δ3D-GLS was −5.6%±4.1. There were no significant differences between treatment groups at baseline. In STUDY, of the 12 participants enrolled, 11 completed a scan at 42 days post FCM treatment. In these, the median age was 47 years, and 91% were women. Median ferritin at referral, and baseline TSAT were 7.4 ug/L (5.3-18.1) and 10.0% (7.8-25.7), respectively.

In Myocardial-IRON, Between-treatment comparisons (FCM vs placebo) showed that lower ferritin values at baseline predicted greater myocardial iron uptake evidenced by a greater decrease in myocardial T1 at 7 and 30 days (Figure 1a). Lower TSAT also identified those with greater drop in myocardial T1 at 7 days but not at 30 days (Figure 1b). Cutoff values for ferritin and TSAT at which myocardial uptake diverged from placebo were <90 ug/L and <18% respectively (Figure 1a and 1b). Lower baseline ferritin was also associated with a greater improvement in 3D-GLS at 30 days (Figure 1c). TSAT did not predict changes in 3D-GLS (Figure 1d).

**Figure 1.**
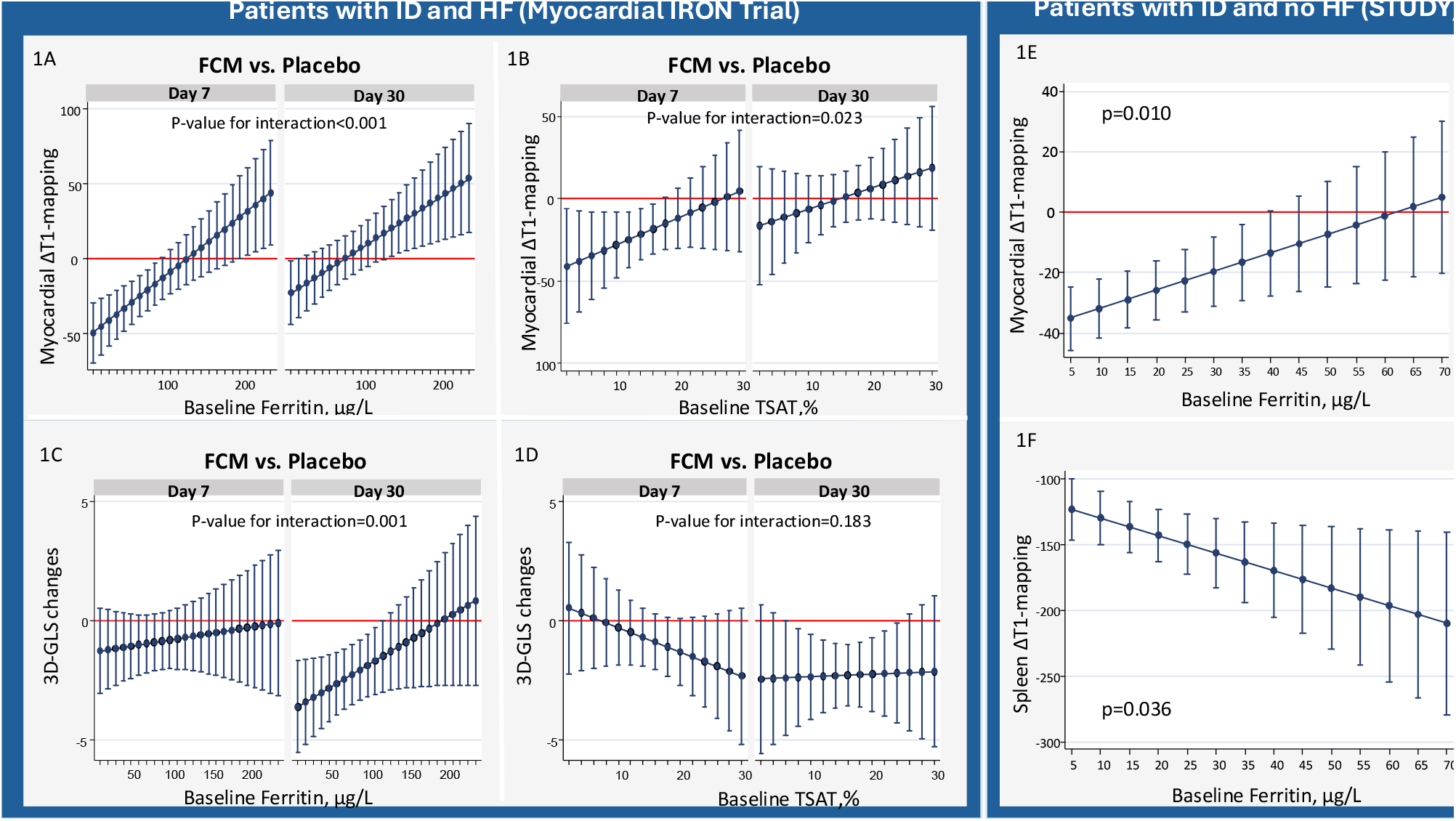
Baseline ferrokinetics and changes in CMR T1-mapping following administration of FCM. 1a. Between treatment-effect (FCM vs placebo) across baseline ferritin in MYOCARDIAL-IRON trial 1b. Between treatment-effect (FCM vs placebo) across baseline TSAT in MYOCARDIAL-IRON trial 1c. Baseline ferritin and changes in 3D-GLS following administration of FCM in MYOCARDIAL-IRON trial 1d. Baseline TSAT and changes in 3D-GLS following administration of FCM in MYOCARDIAL-IRON trial 1e. Baseline ferritin and changes in myocardial T1-mapping (day 42 vs baseline) after FCM therapy in STUDY trial 1f. Baseline ferritin and changes in spleen T1-mapping (day 42 vs baseline) after FCM therapy in STUDY trial

In STUDY, within-group comparison showed that lower baseline ferritin identified patients with a greater myocardial iron uptake as evidenced by greater decrease in myocardial T1 at 42 days (Figure 1e). In contrast, lower baseline ferritin identified those with the least drop in spleen T1 (Figure 1f). Baseline TSAT was not associated with changes in either myocardial or spleen T1 (data not shown).

## CONCLUSION

In patients with or without HF, lower plasma ferritin at baseline predicts greater rise in myocardial iron in the weeks after intravenous iron therapy with FCM. In terms of underlying mechanisms, spleen data from the STUDY trial point towards a plausible hypothesis. As well as raising plasma ferritin, inflammation also promotes iron retention within spleen macrophages. Indeed, hepcidin, the iron regulatory hormone is induced by inflammation and blocks the iron exporter ferroportin on the surface of splenic macrophages (4,5). Our findings that baseline ferritin is a positive predictor of the rise in spleen iron but a negative predictor of the rise in myocardial iron point towards the hypothesis that, in inflamed settings, there is greater trapping of iron within spleen macrophages, which limits iron’s subsequent redistribution to peripheral tissues such as the heart (figure 2). The implication would be that the benefits of IV iron therapy would be limited or short-lived in the setting of functional ID.

**Figure 2.**
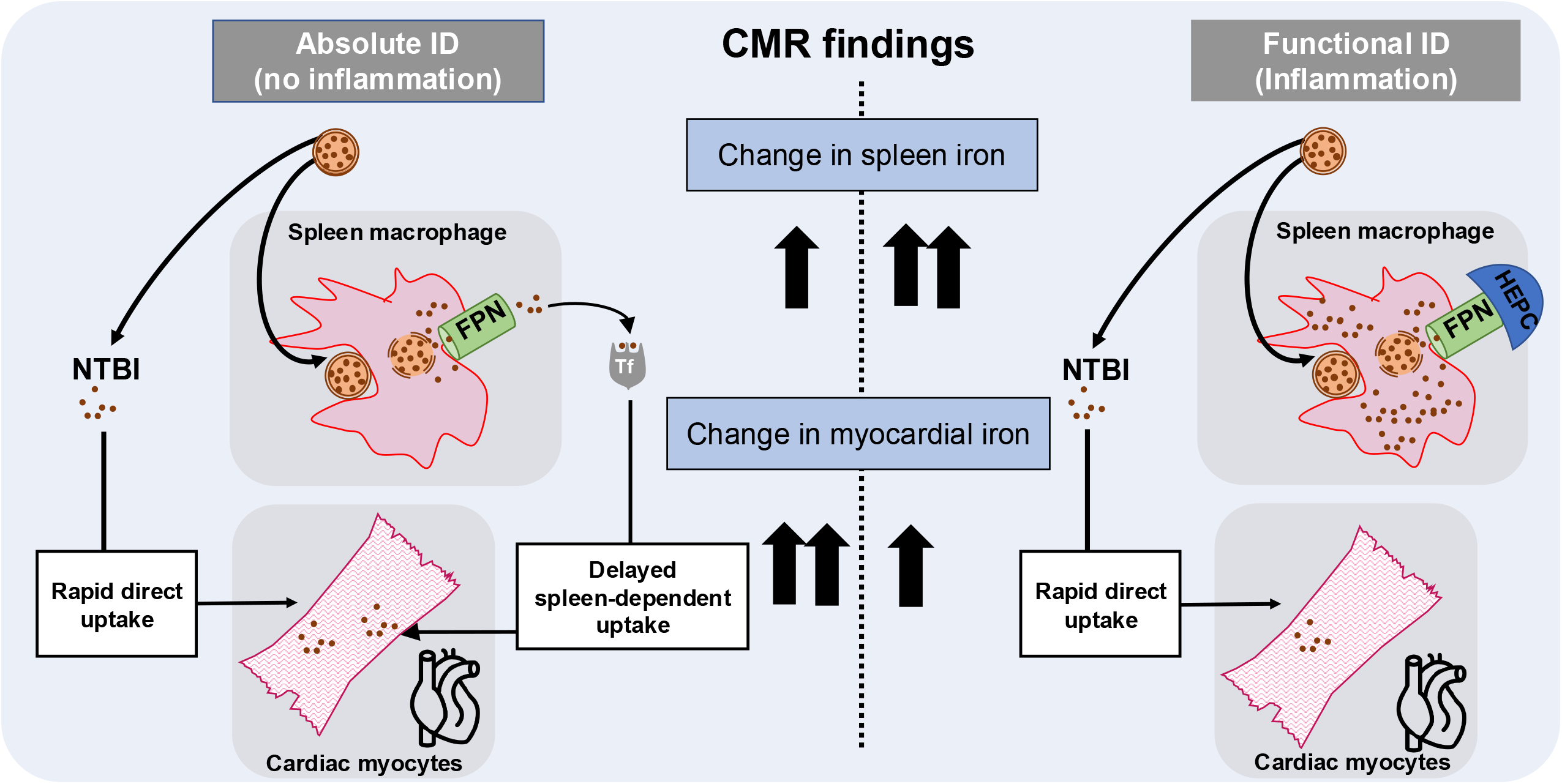
Hypothetical framework for the impact of inflammation on the body’s handling of iron from intravenous iron therapy. Iron carbohydrate complexes such as FCM release some iron directly into the circulation as non-transferrin bound iron (NTBI) which is rapidly and directly taken up by cardiac myocytes, as demonstrated previously by Vera-Aviles M, et al (3). The remainder of iron retained within the carbohydrate complex is taken up by macrophages of the reticuloendothelial system, particularly in the spleen. Here, iron is freed from the carbohydrate complex and exported into the circulation via the iron exporting protein ferroportin (FPN). Upon export into the circulation, iron is loaded onto its chaperone transferrin, and made available for uptake by peripheral tissues including the heart. In the absence of inflammation, low hepcidin levels result in high FPN expression on splenic macrophages, allowing the spleen to redistribute iron from intravenous iron formulations. In the presence of inflammation, high hepcidin (HEPC) levels block FPN on splenic macrophages, causing splenic iron trapping, and limiting the redistribution of iron to peripheral tissues. Thus, in the absence of inflammation, iron from intravenous iron therapies is delivered to the heart both directly (as NTBI) and indirectly via the intermediary of the spleen. However, inflammation blocks the spleen-dependent pathway of iron delivery, thereby limiting the amount of iron taken up into the myocardium.

There is an ongoing debate on what definition of ID identifies HF patients likely to derive the greatest benefit from intravenous iron therapy. Recently, responder analyses of IV iron trials suggested that anemia and functional ID predict greatest benefits (6-9). Our data, on the other hand, suggest that lower ferritin predicts greater myocardial iron repletion, at least in the short-term. One interpretation to reconciles these seemingly divergent results is that myocardial iron repletion is not in fact a mechanism for benefit. An alternative interpretation is that an increase in myocardial iron only benefits those with myocardial iron depletion. Past studies have shown myocardial iron depletion mainly occurs in patients with advanced HF (10), but these patients were not included in the aforementioned IV iron trials. This report generates a plausible hypothesis of how inflammation could limit the benefits of IV iron therapy, and highlights the need for a larger study that formally examines the relationship between myocardial iron repletion and improvements in echocardiographic and clinical outcomes across the ferrokinetic spectrum.

## Data Availability

All data produced in the present study are available upon reasonable request to the authors

## Data Availability

Research data will be made available upon reasonable request to the corresponding author.

